# A Novel Artificial Intelligence Platform to Automate Clinical Consultation Notes and Enhance Diagnostic Efficiency in the Outpatient Clinic: Proposal of a Protocol for a Multi-Center, Multi-Disciplinary, Prospective Randomized Controlled Trial

**DOI:** 10.1101/2023.06.26.23291879

**Authors:** Karanvir Gill, Giovanni Cacciamani, Jamal Nabhani, Joshua Corb, Tom Buchanan, Daniel Park, Virinder Bhardwaj, Onkarjit Marwah, Moses Kim, Deepak Kapoor, Alexander Kutikov, Robert Uzzo, Inderbir Gill

## Abstract

Presented herein is a proposal for a protocol for a multi-center, multi-disciplinary randomized controlled trial (RCT) to evaluate a novel artificial intelligence (AI)-based technology that automates the construction of the clinical consultation note (CCN) and enhances diagnostic assessments in the outpatient clinic setting. This innovative tech-platform automatically generates the CCN and presents it to the provider in advance of the patient consultation, without any work done by the provider. The constructed CCN is presented either in the native electronic health record (EHR) or in a secure web-based application, in a HIPAA-compliant manner. The proposed prospective prospective trial will compare this novel AI/ML technology (NAMT) versus the current standard-of-care (SOC) in the outpatient setting. Outpatient clinic-days will be randomized to either “SOC clinic-day” or the “NAMT clinic-day” based on whether the SOC or the NAMT was used to construct the CCN for all patients seen on that particular clinic-day. Randomized cross-over of each provider between “SOC clinic-day” and “NAMT clinic-day” will result in each provider serving as her/his own internal control. Objective data will be used to compare study endpoints between the SOC and the NAMT. Co-primary endpoints include a) CCN diagnostic accuracy/quality (based on standardized QNOTE metrics); and b) Work-outside-work (WOW) time required by providers to complete clinic-related documentation tasks outside clinic hours (based on EHR meta-data). Secondary endpoints include a) Provider productivity (based on provider “walk-in, walk-out’ time from the consultation room); b) Provider satisfaction (based on the standardized AHRQ EHR End User Survey); and c) Patient satisfaction (based on the standardized Press Ganey/CG-CAHPS survey). To assess generalizability across the health-care spectrum, the study will be conducted in four different types of health-care settings (large academic medical center; non-academic hospital; rural hospital; community private practice); in four different disciplines (cardiology; infectious disease; urology; emergency medicine); using four different EHR systems (Cerner; Epic; AllScripts; MediTech/UroChart). We estimate an aggregate RCT sample size of 150 clinic-days (involving 3,000 total patients; 15-30 providers). This will randomize 75 clinic-days (1,500 patients) to the control SOC arm, and 75 clinic-days (1,500 patients) to the intervention NAMT arm. We will use a two-sided Z-test of difference between proportions with 90% power and two-sided 5% significance level. This RCT is the first to evaluate the efficiency and diagnostic accuracy of pre-constructing CCNs in an automated manner using AI/ML technology, deployed at a large-scale, multi-institutional, multi-disciplinary, multi-EHR level. Results from this study will provide definitive level 1 evidence about the desirability and generalizability of AI-generated automatically constructed CCNs, assessing its potential benefits for providers, patients, and healthcare systems.

## INTRODUCTION

Electronic Health Records (EHR) provide secure, immediate access to patient records, store patient’s clinical information and coding/billing data, and allow patients secure access to their own records, thereby providing comprehensive information at the point-of-care. Following the 2009 Health Information Technology for Economic and Clinical Health (HITECH) Act, EHRs have gained widespread penetration into the U.S. healthcare system, with implementation reaching 96% of U.S. hospitals by 2017 [1, 2].

However, for the providers, EHR usage is labor-intensive by its very nature, with its repetitive data entry now the major contributor to provider burnout nationally [3–8]. In outpatient clinics, providers typically devote ∼50% of their working hours to the EHR, equating to nearly two hours of EHR use for every hour of patient consultation [9, 10]. This substantial time and effort required for EHR data entry detracts from the actual time providers spend interacting with the patient (versus interacting with the computer) during the consultation. This emphasis on EHR data entry results in suboptimal direct communication between providers and patients, detracts from the quality of the consultation, and decreases patient/provider satisfaction, contributing to the current high rates of physician burnout [11]. Additionally, providers often spend considerable amount of personal time after-hours and during weekends to complete EHR-related tasks, termed ‘work-outside-work’ (WOW). Small wonder then that currently up to 70% of U.S. physicians report burnout, directly attributed to EHR usage [11–14]

Provider burnout is linked to various aspects of EHR use, including time spent on EHR, level of organizational support, and system usability [15]. Burnout can decrease the provider’s quality of EHR usage, contributing to diagnostic errors, with implications for patient care [16].

Misalignment of providers’ cognitive processes with the technical aspects of EHR usability and workflow can increase the risk for diagnostic errors [17]. Collectively, these findings highlight the multi-dimensional burdens confronting providers when incorporating EHR into their practice.

During outpatient consultations, providers are required to create a Clinical Consultation Note (CCN) for each patient, which consolidates the patient’s medical history, prior test data, and treatment plans in a concise and structured format. Constructing the CCN in the outpatient clinic takes on average 16 minutes of EHR usage per patient [12]. Thoroughness of the CCN drives ICD clinical coding [18], and high CCN quality is also associated with improved physician performance and patient outcomes [19]. Objective, standardized tools to assess CCN diagnostic efficiency and quality are available [20, 21], and EHR metadata can help evaluate the individual physician effort invested into creating this document [9, 10].

Emerging artificial intelligence (AI) and machine learning (ML) technologies present robust new opportunities for healthcare innovation [22]. Given that a significant portion of CCN creation involves repetitive, scribe-like work, it can be mitigated with technology. Efforts are underway to use speech-to-text technology and AI to automate CCN creation during clinic visits [9, 10]. More recently, a novel AI/ML and logic-based technology platform has been developed that automates the construction of a detailed preliminary CCN before the actual consultation, without any work on the part of the provider.

Herein, we intend to perform a prospective randomized controlled trial (RCT) to compare this novel technology platform with current standard-of-care practices in the outpatient clinic setting. We seek to evaluate the benefits and efficiencies that may accrue to providers and patients alike from using AI to automate the creation of the CCN. Using a large multi-institutional, multi-disciplinary, multi-EHR cohort, our RCT will compare the novel technology (wherein the CCN is constructed automatically using AI before the actual consultation) vis-s-vis the current standard-of-care practice (wherein the CCN is constructed manually during the actual consultation). The generated data will assess CCN diagnostic efficiency and quality, provider ‘work-outside-work’ effort, provider productivity, provider satisfaction, and patient satisfaction.

## NOVEL TECHNOLOGY

The novel AI technology is comprised of a proprietary logic algorithm-based, symptom-tree Active Sheet that extracts subjective patient data directly from the patient and analyzes and reformats it in a structured manner. In parallel, the AI/ML algorithm extracts relevant, unstructured, objective data from outside records/documents. Proprietary, embedded AI/ML technology generates national guidelines-based recommendations for diagnostic testing in a patient-specific manner. This preliminary CCN is automatically presented to the provider, in advance of the patient’s visit, either within the institution’s EHR or in a secure web-based App, in a HIPAA-compliant manner. During the actual consultation, the provider re-confirms the clinical history with the patient, performs the physical exam, and finalizes the CCN, completing the patient consultation. The platform’s combination of AI/ML algorithms, Active Sheet logic algorithms, and Optical Character Recognition (OCR) technology makes it adaptable to various types of data inputs. Its capability of EHR-integrated and web-based deployment, as well as usage via laptops and cell phones, also ensures high scalability, allowing implementation in an institution with or without an existing EHR system, and for patients with or without laptop computers.

## METHODS

Our study protocol follows the Standard Protocol Items Recommendations for Interventional Trials-Artificial Intelligence (SPIRIT-AI) Guidelines [23]. The current manuscript is only a proposal for the protocol for this study; IRB approval and clinicaltrials.gov notification will precede the actual trial initiation and consenting/enrollment of any clinical patients.

### Study Settings

This is a prospective multi-institutional, multi-disciplinary, multi-EHR randomized trial. To assess generalizability across the health-care landscape, the RCT will be conducted concomitantly across four different types of healthcare settings: large academic medical center, non-academic hospital, rural hospital, and community private practices. Participating centers include the University of Southern California and Fox Chase Cancer Center (large academic medical centers); Sierra View Medical Center, Verdugo Hills Hospital (non-academic, rural hospitals); and Solaris Urology and Glendale Cardiology (community private practices). Four specialties will be assessed, including two medical specialties (cardiology, infectious disease), a surgical specialty (urology), and an urgent care specialty (emergency medicine). Four different types of EHRs will be assessed, including Cerner, Epic, AllScripts, and MediTech.

This prospective trial will randomize the novel AI/ML technology (NAMT) versus the current standard-of-care (SOC) in the outpatient setting. Randomization will be done per outpatient clinic-days based on method of constructing the CCN. As such, randomization to either the “SOC clinic-day” or the “NAMT clinic-day”, will be based on whether the SOC or the NAMT was used to construct the CCN for all patients seen by that provider in that clinic on that day.

Randomized cross-over of each provider between “SOC clinic-day” and “NAMT clinic-day” will allow each provider to serve as his/her own internal control. To provide a balanced and valid comparison, we will use similar clinic workflow methodology for collecting outside records/data in each arm. Thus, medical records clerks and/or nurses would collect any available outside patient records/data in both arms per existing, legacy data collection methods at each participating site. In the “SOC clinic-day” arm, the medical records clerk/nurse would upload patient records to the native EHR platform, and then the CCN would be constructed manually per current standard practice. In the “NAMT clinic-day” arm, the medical records clerk/nurse would upload these records to the NAMT platform (typically housed within the native EHR, thus maintaining similarity in data/records collection methods), but the CCN would be constructed automatically by the NAMT platform. Thus, we will use the same data entry mechanism in each arm to ensure consistency and enhance validity of results. Similar objective endpoint data will be obtained for each provider and patient in the control “SOC clinic-day” arm and the experimental “NAMT clinic-day” arm (Figure 1 – CONSORT-AI Flowchart [24]).

**Figure 1:**
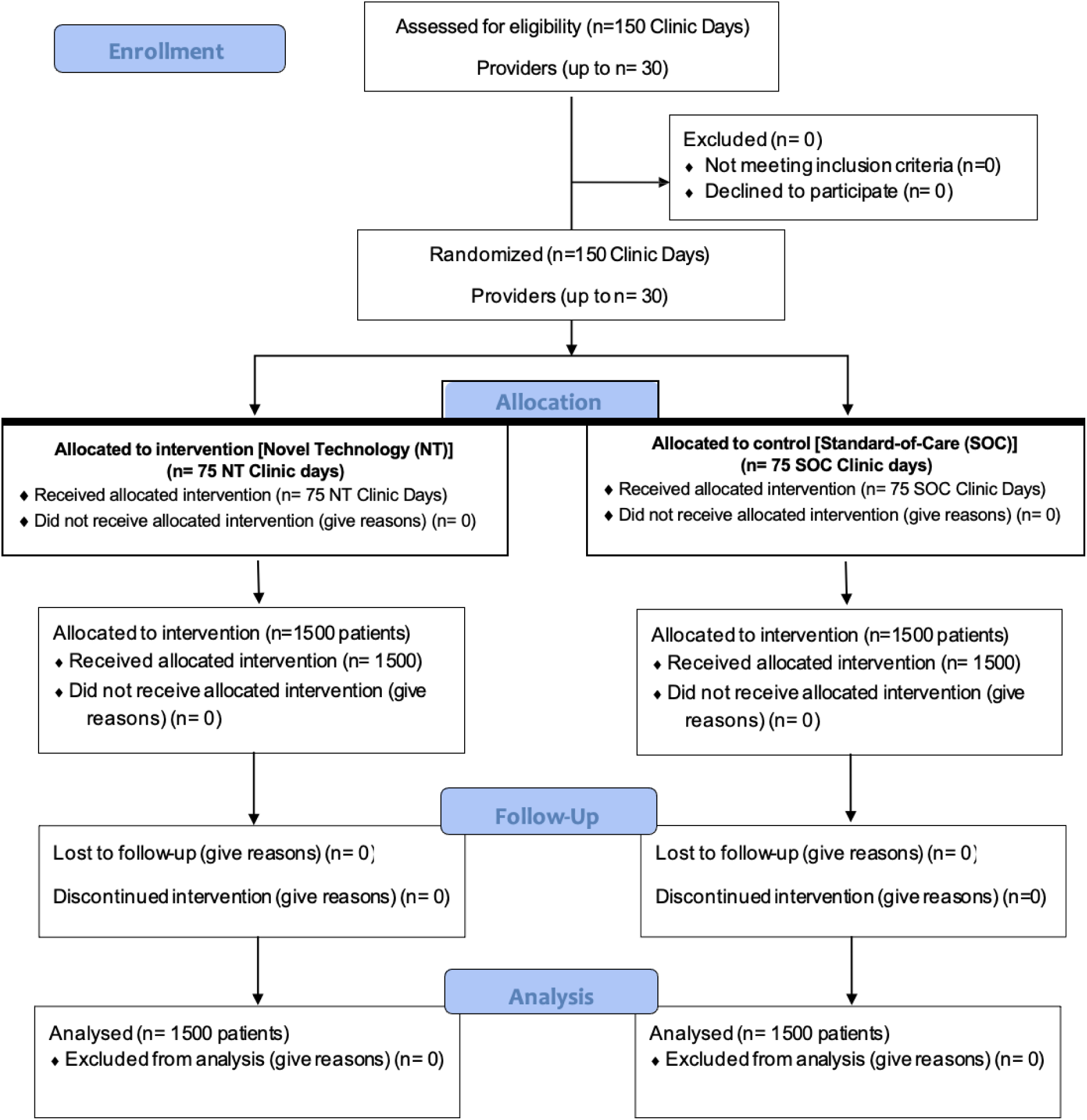
CONSORT Flow Diagram (Ideal Scenario)

Study endpoints will compare the SOC vs the NAMT based on objective data. Co-primary endpoints are: a) CCN quality and diagnostic accuracy (based on standardized QNOTE metrics [14]); and b) Work-outside-work (WOW) time required by providers to complete clinic-related documentation tasks outside normal clinic hours (based on EHR meta-data). Secondary endpoints include: a) Provider satisfaction (based on standardized AHRQ EHR End User Survey); b) Patient satisfaction (based on standardized Press Ganey/CG-CAHPS survey (www.pressganey.com)); and c) Provider productivity (based on provider “walk-in, walk-out’ time from the individual consultation room) and imputed financial data. In total, 15-30 providers will participate. Provider is defined as a licensed medical provider who is constructing the CCN in their daily practice currently, including practicing physicians, mid-level practitioners (physician assistant, nurse practitioner), or trainees (post-graduate fellow or resident). Scribes are not designated as a provider. At the time of scheduling the clinic appointment, patients will be informed that, on their specific clinic-day, their provider is doing all consultations using the specific methodology (SOC or NAMT), as dictated by the randomization. At study onset, all participating providers will attest that they have equipoise between the SOC and NAMT approaches.

### Randomization: Rationale

Our objective is to evaluate the merits/demerits of the novel AI/ML technology for the automated construction of CCN vis-a-vis the standard-of-care (SOC). Elements that could potentially be randomized include patients, providers, or the intervention itself. However, practically speaking, all of these would impact daily clinic workflow. For instance, if we were to randomize patients, then using both the SOC and NAMT within the same clinic day would be highly disruptive to clinic workflow, and challenging, if not prohibitive, to implement from a practical perspective. If we randomized providers, it would be problematic to randomize the AI/ML technology, and vice-versa. Also, providers with different practice styles conducting one or the other intervention would not control for inter-personal disparities. However, randomization of the entire clinic-day to either “SOC clinic-day” or “NAMT clinic-day” for all patients seen that day, and then equally alternating each provider between “SOC clinic-day” and “NAMT clinic-day” would satisfy study objectives and allow each provider to serve as her/his own internal control. Providers would be a heterogeneous group as regards gender, race, and age. This study design is pragmatic and would be minimally disruptive to daily clinic workflow or provider productivity. To minimize provider bias, at enrollment, all consented providers would attest to equipoise between SOC and NAMT.

### Eligibility Criteria

Inclusion and exclusion criteria are reported separately for providers and patients.

#### a) Providers

i. Inclusion criteria: Licensed medical providers currently engaged in patient consultations in the outpatient clinic, including practicing physicians; mid-level practitioners (physician assistant, nurse practitioner); or trainee (post-graduate fellow or resident). Use of scribes by providers is allowed.
ii. Exclusion Criteria: Licensed medical providers who are currently not engaged in patient consultations in the outpatient clinic.

#### b) Patients

i. Inclusion criteria: Outpatient clinic patients at least 18 years of age or older being seen in the cardiology, infectious disease, urology, and emergency medicine outpatient settings.
ii. Exclusion criteria: Patients younger than 18 years of age.

### Intervention

Licensed, actively practicing providers who satisfy inclusion criteria are recruited after written consent. Their upcoming clinic days are randomized to either “SOC clinic-day” or “NAMT clinic-day”, wherein CCNs for all patients seen on those respective days are constructed using either the standard-of-care practice or the novel AI/ML technology (Figure 1). Clinic days will be randomized 1-2 weeks prior to the actual date of that clinic. As such, all patients seen on the clinic day randomized to the “SOC clinic-day” will have their CCN constructed per that provider’s customary, standard-of-care practice. Patients seen on the clinic day randomized to the “NAMT clinic-day” will have their CCN constructed 2 weeks to 1 day prior to the clinic visit using the novel AI/ML technology. The automatically constructed CCN will be made available to the provider in advance of the clinic day, either integrated within their native EHR or in the secure, free-standing web-based App. After pre-viewing the CCN, the provider will proceed to perform the clinical consultation. Provider “walk-in, walk-out” time from the consultation room will be recorded by a third-party observer using a stopwatch. After completing the consultation, the patient will be invited to complete the adapted Press Ganey/CG-CAHPS patient satisfaction questionnaire; the patient will provide oral or written informed consent before filling out the de-identified survey instrument. This survey questionnaire has been adapted to fit our study-related objectives, including questions on patient age, gender, level of education, race/ethnicity, provider attentiveness to the patient, provider spending time doing computer work versus interacting with the patient, and provider’s prior knowledge about the patient’s clinical condition before entering the patient’s consultation room. The questionnaire then provides the patient a final opportunity to record an overarching ‘thumbs-up’ or ‘thumbs-down’ about their clinic experience. Interventions for each group are provided below with sufficient details to allow replication, including method and timing of administration.

#### a) Study Intervention for providers

Providers will be invited to participate in the study and informed consent obtained. Providers and their teams will be trained on the novel technology and related clinical workflow over a period of 1-3 clinic-days (“Wash-in” period) to make them knowledgeable about the novel technology platform.

Outpatient clinic days will be randomized into one of 2 arms, “SOC clinic-day” and “NAMT clinic-day”. (Details in Figure 1)

i. On “SOC clinic-day”: CCNs for all patients seen on this day will be constructed per the usual, standard-of-care practice of that provider. Prior to the clinic day, the medical records clerk/nurse would upload any outside patient records/data to the native EHR platform per current standard practice, and then the CCN would be constructed manually.
ii. On “NAMT clinic-day”: CCNs for all patients seen on this day will be pre-constructed using the novel AI/ML technology. At the time of being given the clinic appointment, patients will be sent the electronic link and advised to do two things from their home - ‘the earlier, the better’ - prior to coming to the clinic: a) complete the Active Sheet (AS): when completing the AS, the patient may solicit help from family/friends, or from remote licensed provider extender (PA/NP) or referring provider staff; and b) upload any lab/radiology test results: the medical records clerk/nurse would help the patient upload outside records/data to the NAMT platform (typically housed within the native EHR platform). After these two actions – completing the AS; uploading the lab/radiology test results - are done, the novel AI software automatically constructs the CCN and delivers it to the provider, either in their native EHR or in the secure, free-standing web-app. This action occurs automatically in the background invisible to the provider/staff, in advance of the patient’s visit. The patient then comes to the clinic on the scheduled appointment date, the provider reviews the pre-constructed CCN and proceeds with the consultation.

#### b) Study Intervention for patients

Immediately upon completion of their consultation, patients will be invited to fill out the adapted Press-Ganey/CG-CAHPS survey in a de-identified fashion to assess patient satisfaction. Patients who agree to fill out the survey will be asked to first provide written informed consent for inclusion in the study. Upon completion of the survey, the patient departs the clinic area. Patient participation will be deemed completed upon survey completion.

### Outcomes and Measurements

Table 1 summarizes the a) co-primary and b) secondary outcomes of interest. Details regarding the outcome measurements are reported in the supplementary materials.

**Table 1.**

| OBJECTIVES | ENDPOINTS | JUSTIFICATION FOR ENDPOINTS | PUTATIVE MECHANISMS OF ACTION |
| --- | --- | --- | --- |
| **CO-PRIMARY** |  |  |  |
| 1. Quality/Diagnostic Accuracy of Clinical Consultation Note (CCN) | 1. QNOTE: to assess CCN quality/diagnostic accuracy (*Objective data*) | 1. The quality/accuracy of the automated CCN should be, at a minimum, equivalent to the standard-of-care | 1. QNOTE: Blinded, objective third-party assessment of CCN quality/accuracy |
| 2. Efficiency of CCN construction | 2. Objective Meta-data: from EMR (*Objective data*) | 2. Extra time (beyond typical clinic hours) spent by provider to complete CCN should decrease | 2. Work-outside-Work (WOW) time/provider: Objective per-patient EHR meta-data from IT team |
| **SECONDARY** |  |  |  |
| 3. Provider satisfaction | 3. AHRQ EHR End User Survey (*Objective data*) | 3. Automating the CCN should decrease provider workload & improve satisfaction | 3. Providers to fill out adapted AHRQ EHR End User Survey per new patient consult or per clinic-day |
| 4. Patient satisfaction | 4. Press Ganey/CG-CAHPS survey (*Objective data*) | 4. Automating the CCN should allow provider to spend more time interacting with the patient (rather than with the EHR), improving patient satisfaction | 4. Patients to fill out adapted Press Ganey/CG-CAHPS survey at end of the consult |
| 5. Provider productivity | 5 Consultation time savings (*Objective data*) | 5. Consultation times should, at a minimum, be similar to standard-of-care, upon automating the CCN | 5. Physician “walk-in walk-out” time from consult room, measured by stopwatch |

#### a) Co-Primary Outcomes

Our co-primary objectives are (a) delivering a CCN of high-quality and diagnostic accuracy without any effort on the part of the provider, and (b) reducing the provider time for doing work-outside-work (WOW) to complete clinic-related documentation activities performed outside of the typical 8 am to 4 pm clinic schedule. Corresponding co-primary endpoints are the objective data metrics to assess CCN quality and diagnostic accuracy (using the standardized QNOTE methodology), and provider WOW time (using EHR meta-data). The rationale behind these endpoints is rooted in the central tenet of this automated technology, which is to create an automated CCN that matches or exceeds the current, manually created, standard-of-care CCN, while minimizing workload for the healthcare provider. Furthermore, we aspire to decrease the time spent by providers working on clinic notes outside regular work hours. To ascertain these endpoints, we utilize QNOTE to measure CCN quality and diagnostic accuracy using objective metrics; provider’s WOW time is determined by analyzing the objective EHR metadata for that clinic-day, provided by the institutional IT team. The putative mechanisms of action underpinning these objectives lie in streamlining clinical processes, thus enhancing the quality of patient care and reducing diagnostic errors.

#### b) Secondary Outcomes

The secondary objectives of the study assess provider satisfaction, patient satisfaction, and physician productivity. Provider satisfaction will be assessed using an adapted AHRQ Electronic Health Record End User Survey. We hypothesize that automating the CCN will reduce the workload of physicians, and consequently improve satisfaction and decrease burnout rates. At a minimum at the end of each clinic day, providers will fill out the AHRQ EHR End User Survey to capture their clinic-day experiences and responses.

Patient satisfaction, another critical component of healthcare delivery, will be evaluated using an adapted Press Ganey/CG-CAHPS survey. We hypothesize that if providers spend more time directly interacting with patients, instead of interacting with and entering data into the EHR, patients will experience higher satisfaction. As such, consented patients will be requested to complete the adapted Press Ganey/CG-CAHPS survey immediately after their consultation to measure their level of satisfaction. Finally, physician productivity, particularly in terms of time savings, which can lead to increased patient throughput and financial return-on-investment (ROI), will be documented. We believe that automating the CCN might lead to decreased, yet higher quality, consultation times, thereby improving physician productivity. To measure this, the physician’s “walk-in, walk-out” time from each consult room will be recorded with a stopwatch. This objective, real-time measurement will assess the impact of the intervention on physician productivity. Financial ROI will be imputed.

### Sample Size

Given the novelty of our AI algorithm, prior randomized data do not exist. Our sample size will be designed to detect a minimum of 30% improvement in the CCN quality/diagnostic accuracy, and a 30% reduction in the duration of WOW, our 2 co-primary endpoints. We estimate the total sample size to be 150 clinic-days, i.e., 75 “SOC-days” and 75 “NAMT-days”. Conservatively assuming 20 patients seen per clinic-day, we will accrue at least 3,000 total patients - 1,500 in the control “SOC-day” arm, and 1,500 in the intervention “NAMT-day” arm. Clinic-days will be divided between the 4 specialties, leading to 30-40 clinic-days per specialty. We estimate a total of 15-30 study providers, resulting in 5-10 clinic-days per provider. No single specialty will accrue more than 30% of the aggregate clinic-days. We will use a two-sided Z-test of the difference between proportions with 90% power and two-sided 5% significance level. This sample size exceeds minimum requirements and will provide robust data. Study data will be collected by trained personnel, physically located at each clinical site. The validated QNOTE scoring system will assesses quality and diagnostic accuracy of de-identified CCNs uploaded to an online repository for scoring by independent, blinded raters. Data collection process will be integrated into clinic workflows, preserving the user experience while introducing the new AI/ML tool.

### Recruitment

Providers will be recruited from the participating institutions. We anticipate between 15-30 providers to participate in this RCT. This would provide a heterogeneous group of providers as regards gender, race, and age, with each provider participating in 5-10 study clinic-days. Randomized alternating cross-over of providers will result in each provider serving as her/his own internal control.Patients will be identified exclusively based on the participating providers’ clinic schedules. Patients will be recruited and consented to the study immediately after they finish their clinical consultation with the provider, and before they are administered the Press Ganey/CG-CAHPS survey instrument. We intend to recruit over 3000 patients over a one-year period. No incentives will be provided to either physicians or patients for participating in this RCT.

### Screening Failure

In case a provider has been already enrolled but wishes to drop out of the study within the first 5 clinic-days, he/she will be replaced with another physician from that specialty. Any provider who has already completed the first 5 clinic-days in the study will not be replaced by another provider; their data will be used as is with censoring at study drop-out. If a patient chooses to not fill out the Press Ganey/CG-CAHPS satisfaction survey, it will be recorded as such, and their survey data will be excluded from study calculations.

### Discontinuation of intervention

After initially agreeing to participate, if a provider decides to discontinue in the study, she/he must express this desire prior to 5 clinic-days after enrolling in the study; in this case their data over those 5 clinic-days will be used, and they will be replaced by another physician. After 5 clinic-days, they will be considered to have been part of the trial and while their data will be included for analyses, they will not be replaced by another physician. Participation in this study is voluntary. If a patient decides to not provide informed consent at the time of answering the Press Ganey/CG-CAHPS questionnaire, their survey data will not be included in the study. If a patient decides to not participate in the study after initially agreeing, they must express this desire prior to completing their Press Ganey/CG-CAHPS questionnaire and leaving the clinic. The sole criterion for patient participation in the study is patient willingness. Participation in this study is voluntary.

### Statistical Hypothesis and Analysis

#### a) Statistical Hypothesis

i. Primary Efficacy Endpoint(s): We hypothesize that the CCN generated by the novel technology will be superior to the SOC as regards CCN quality and diagnostic accuracy (as measured by QNOTE methodology), and CCN efficiency (WOW time as measured by EHR meta-data). Our null hypothesis is that there will be no difference in the quality, diagnostic accuracy, and efficiency of the CCN derived from the novel technology compared to standard-of-care.
ii. Secondary Efficacy Endpoint(s): We hypothesize that the CCN generated by the novel AI/ML technology will be superior to the SOC as regards provider satisfaction (adapted AHRQ Electronic Health Record End User Survey); patient satisfaction (adapted Press Ganey/CG-CAHPS survey); and physician productivity (“walk-in, walk-out” consult time).

Our null hypothesis is that outcomes will be no difference in the above-mentioned three metrics for CCNs generated by the novel AI/ML technology versus standard-of-care.

#### b) Statistical analysis

Analysis of the primary endpoint will assess mean data for CCN quality/accuracy and CCN efficiency to demonstrate superiority of the NT compared to SOC.

i. Quality of CCN will be assessed by the adapted QNOTE instrument, a validated methodology that utilizes 15 clinical elements and 7 evaluative components (as detailed in supplementary materials). Each CCN will be de-identified by redacting patient/physician identifiers prior to assessment and uploaded to an online repository for QNOTE scoring by three blinded, third-party raters. Each patient will be assigned a unique identifier number to ensure confidentiality and privacy. Depending on quality of each CCN, a score between 0 (minimum) to 1500 (maximum) will be assigned by independent raters. The average of the ratings from the 3 independent raters for each CCN will denote the composite QNOTE score for that CCN, which will be the basis for statistical analysis comparing CCNs created on “SOC clinic-day” versus “NT clinic-day”.
ii. Work-outside-Work (WOW) time will be objectively assessed from EHR meta-data per provider (as detailed in supplementary materials).

Analysis of the secondary endpoints will calculate mean data to assess outcomes of the NT compared to SOC for the following three aspects:

i. Provider satisfaction (adapted AHRQ Electronic Health Record End User Survey): dichotomous positive versus negative Likert scale answers will be compared for each question and in the aggregate.
ii. Patient satisfaction (adapted Press Ganey/CG-CAHPS survey): dichotomous positive versus negative Likert scale answers will be compared for each question and in the aggregate.
iii. Provider productivity (Total consultation time): physician “walk in-walk out” time from the consult room at each patient consultation will be measured by the study coordinator using a stopwatch.

The sample size will be designed to detect a minimum of 30% improvement in the CCN quality/diagnostic accuracy, and a 30% reduction in the duration of WOW, our 2 co-primary endpoints. Analysis of primary and secondary objectives will proceed as follows. Missing data will be dealt with by imputation, and potential outliers will be scrutinized by the Principal Investigator. For each question, Likert scale responses will be analyzed, allowing comparisons between the novel technology and the standard-of-care. Answers will be divided into positive and negative categories per the “top box method,” which is standard for assessing CG-CAHPS data. Likert scale responses will be dichotomized, with scores of 4 (agree) and 5 (strongly agree) representing agreement (positive) and scores of 1 (strongly disagree), 2 (disagree), or 3 (neutral) indicating disagreement (negative). Screening for outliers will be conducted based on the absolute agreement of each individual question and distribution of responses. Continuous variables will be represented as mean and standard deviation (SD), and categorical variables will be presented as median and interquartile range (IQR). For the univariate analysis, Kruskal-Wallis, chi-squared (X2), and Fisher’s exact tests will be utilized to compare continuous and categorical variables as appropriate. A 95% confidence interval and a two-tailed test with p < .05 will be taken as statistically significant.

Upon completion of data collection, each question will be analyzed using Cochran-Mantel-Haenszel tests, stratified by provider to eliminate potential individual provider bias from the results. P-values will be determined using Chi-Square tests and will be deemed significant at less than 0.05. For the assessment of patient satisfaction (utilizing the adapted Press Ganey/CG-CAHPS survey with additional queries), each question will be stratified by provider to remove potential individual bias and analyzed using Cochran-Mantel-Haenszel tests.

Considering the study’s novelty and the absence of prior randomized data, an interim analysis will be conducted. Calculations will be performed using an α level of 0.05 and a power of 90%. Based on the trends in the data from the first 50 patients, sample sizes will be calculated for each question, assuming the variables to be dichotomous. After the complete data collection, each question will be analyzed using Cochran-Mantel-Haenszel tests, stratified by providers to remove any individual provider bias. P-values will be determined using Chi-Square tests, and a value of less than 0.05 will be considered significant.

## CONCLUSION

Herein we propose the protocol of the first prospective randomized controlled trial of a large-scale, multi-institutional, multi-disciplinary cohort to evaluate the practicality and efficiency of using AI/ML technology to pre-construct the CCN automatically before the actual patient-provider consultation. The study will compare this AI/ML framework vis-a-vis the standard-of-care, wherein the CCN is generated manually during the consultation encounter. Data generated by this study will provide insights into the desirability and generalizability of AI-generated automated clinical consultation notes across the healthcare spectrum, and its potential benefits to providers, patients, and healthcare institutions.

## Data Availability

No data described in the present study

